# Estimating the burden of malaria and soil-transmitted helminth co-infection in sub-Saharan Africa: a geospatial study

**DOI:** 10.1101/2022.03.14.22272330

**Authors:** Muhammed O. Afolabi, Adekola Adebiyi, Jorge Cano, Benn Sartorius, Brian Greenwood, Olatunji Johnson, Oghenebrume Wariri

## Abstract

**Background:** Limited understanding exists about the interactions between malaria and soil-transmitted helminths (STH), their potential geographical overlap and the factors driving it. This study characterised the geographical and co-clustered distribution patterns of malaria and STH infections among vulnerable populations in sub-Saharan Africa (SSA).

**Methodology/Principal findings:** We obtained continuous estimates of malaria prevalence from the Malaria Atlas Project and STH prevalence surveys from the WHO-driven Expanded Special Project for the Elimination of NTDs (ESPEN) covering 2000-2018 and used spatial autocorrelation methods to identify statistically significant clusters for both diseases across SSA. We used the inverse distance weighted kriging (interpolation) methods to estimate STH prevalence. We calculated the population-weighted prevalence of malaria and STH co-infection, and used the bivariate local indicator of spatial association (LISA analysis) to explore potential co-clustering of both diseases at the implementation unit levels.

Our analysis shows spatial variations in the estimates of the prevalence of *Plasmodium falciparum*-STH co-infections and identified hotspots across many countries in SSA with inter-and intra-country variations. High *P. falciparum* and high hookworm co-infections were more prevalent in West and Central Africa, whereas high *P. falciparum*, high *Ascaris lumbricoides*, high *P. falciparum*, and high *Trichuris trichiura* co-infections were more predominant in Central Africa, compared to other sub-regions in SSA.

**Conclusions/Significance:** Wide spatial heterogeneity exists in the prevalence of malaria and STH co-infection within the regions and within countries in SSA. The geographical overlap and spatial co-existence of malaria and STH could be exploited to achieve effective control and elimination agendas through the integration of the vertical programmes designed for malaria and STH into a more comprehensive and sustainable community-based paradigm.

**Author Summary:** Malaria and worms frequently co-exist together among children living in the poorest countries of the world, but little is known about the specific locations of the combined infections involving the two major parasitic diseases and how they interact and change over the years.

We used open access data collected by two public registries, that is, the Malaria Atlas Project and Expanded Special Project for the Elimination of NTDs, to understand the overlap of the two diseases in different parts of Africa, where their burden are more predominant.

We found significant differences in the distributions of the combined diseases across different parts of Africa, with large concentrations identified in Central and West Africa. For example, double infections with malaria and hookworm were more common in West and Central Africa, whereas malaria and roundworm, and malaria and whipworm were predominantly found in Central Africa. A large collection of the dual infections was also found in some localities within the countries which appeared to have low burden of the two diseases.

These findings provide a useful insight into the areas which could be serving as a reservoir to propagating the transmission of the two diseases. The results of this study could also be used to develop and implement integrated control programmes for malaria and worms, and this could help to achieve the WHO NTD roadmap to ending the neglect to attain Sustainable Development Goals by 2030.

## Introduction

Due to environmental and host factors which favour transmission of multiple parasitic infections, malaria and soil-transmitted helminths (STH), including *Ascaris lumbricoides, Trichuris trichiura*, and hookworms, co-exist in many parts of the world, predominantly in sub-Saharan Africa (SSA) [1], where they are most prevalent. An estimated 241 million cases of malaria were reported in 2021 in 85 malaria endemic countries, 29 of which accounted for 96% of malaria cases globally, and six SSA countries (Nigeria, Democratic Republic of the Congo, Uganda, Mozambique, Angola and Burkina Faso) carried more than half of this burden. [2]

Similarly, a disproportionately high burden of STH has been reported in many countries in SSA with large concentrations of moderate-to-heavy intensity infections in Nigeria, Democratic Republic of the Congo, Ethiopia, Cameroon, Angola, Mozambique, Madagascar, Equatorial Guinea, and Gabon. [3]

In these malaria-STH co-endemic countries, estimating the national burden of the co-infection poses a public health challenge because national surveillance systems are often sub-optimal.[4] For example, deworming through mass drug administration (MDA) of anthelminthics is widely regarded as a cost-effective strategy. However, the impact of these MDA programmes on the changing prevalence and intensity of STH in children, is often hindered by a lack of comprehensive baseline data before MDA initiation, because of financial challenges or as a result of the administration of the drugs via undocumented channels.[3] Nevertheless, recent WHO reports [1, 4] indicate that considerable progress has been made in reducing the overall burden of malaria and STH in many countries including some in SSA. To sustain the gains of past decades and move the control efforts towards elimination, WHO has recommended a paradigm shift from disease-specific to an integrated approach.[4]

Empirical studies conducted across co-endemic countries have shown that variations in the prevalence of malaria and STH co-infections differed significantly across geographic locations.[5-9] Limited understanding exists about the spatial distribution of these co-infections in vulnerable populations. Majority of the published studies targeting malaria-STH co-infection have focused on describing the transmission and burden among affected populations, [5, 6, 9, 10] and have paid little attention to their concurrent spatial distribution and causes. Previous systematic reviews have also reported an over-estimation of the relationship between malaria and helminths making it difficult to establish conclusively the burden of malaria-STH dual infections. [11, 12] Consistent with findings of these reviews, a recently conducted systemic review and meta-analysis of 55 studies which enrolled 37,559 children across low and middle-income countries (LMIC) found a wide variation in the prevalence of malaria-helminth co-infections, ranging from 7-76% across LMICs.[13] These findings may be due to the low sensitivity of the diagnostic methods employed for the detection of malaria-helminth co-infections in the primary studies included in the systematic review.

The WHO road map for the control and elimination of NTDs for the 2021-2030 period defines STH elimination as a public health problem (EPHP) when there is a prevalence of less than 2% of moderate-to-heavy intensity infections.[14] However, as STH control accelerates and new detection tools are developed, a significant number of people with clinical features outside high intensity infection hotspots are now being seen. [15]

Reliable estimates of malaria-STH morbidity hotspots are needed to comply with the NTD roadmap goal of intensifying cross-cutting approaches which integrate STH control with common delivery platforms for diseases that share similar epidemiology such as malaria. Also, empirical evidence shows that obtaining reliable estimates of the burden of STH-malaria co-infection would help address the critical gaps in planning and implementation of integration of STH control strategies with other interventions. [14]

We undertook a geospatial analysis on the datasets describing the prevalence of malaria and STH infections among adults and children aged 5-14 years, obtained from the geospatial open-access data generated from the Malaria Atlas Project (MAP)[16] and from the WHO-driven Expanded Special Project to Eliminate NTDs (ESPEN) [17], to characterise the geographical and co-clustered distribution patterns for malaria and STH co-infections among vulnerable populations in SSA.

## Methods

### Study design

We analysed the spatial relationships between STH and malaria using spatial autocorrelation methods to identify statistically significant clusters over different regions in endemic countries in SSA. Based on the classifications of the United Nations Geoscheme for Africa[18], we categorised SSA into west, east, central and southern African sub-regions. These include 46 of Africa’s 54 countries and territories that are fully or partially south of the Sahara, excluding Algeria, Djibouti, Egypt, Libya, Morocco, Somalia, Sudan and Tunisia.

We obtained datasets on the prevalence of *Plasmodium falciparum* from the MAP database (last accessed September 2, 2021). MAP is a WHO collaborating platform for geospatial disease modelling which was created for the purpose of determining spatial limits, prevalence and endemicity o*f P. falciparum and P. vivax*. MAP provides a database to obtain data on the estimated prevalence of *P. falciparum and P. vivax* at 5km resolution for each year from 2000 onwards. We focused on *P. falciparum* in this study because it is the predominant causative agent for more than 95% of malaria cases in SSA[19]. We also obtained datasets on the prevalence of STH (*A. lumbricoides, T. trichiura* and hookworms) from the WHO-driven ESPEN portal. ESPEN portal contains survey datasets on NTDs in Africa. Survey datasets from both MAP and ESPEN were confined to surveys carried out from 2000 to 2018.

The *P. falciparum* prevalence data obtained from MAP were provided in a raster format for each year covering the entire African continent. To make them usable, we combined the data into one raster file using a mosaic tool. The description of the methodology used for generating the predicted prevalence is available at https://malariaatlas.org/api-docs/

The prevalence datasets on STH (*A. lumbricoides, T.trichiura*, and hookworms) extracted from the ESPEN portal were geolocated, that is, the data contains the geographic coordinates of the survey locations which can be either a school or village location. The data consist of 25,274 data points from 2000 to 2018, of which 817 (3.2%) were without spatial reference. Of the remaining 24,457 (96.8%) data points with spatial reference, 6,229 data points had prevalence rates above zero for *T. Trichiura*, 12,945 data points had prevalence rates above zero for hookworm and 9,728 for *A.lumbricoides*. Incomplete data (those without spatial reference and explicit prevalence rates) were excluded from the analyses.

## Statistical analysis

### Spatial auto-correlation

We used Moran’s I statistics as the indicator of spatial auto-correlation. We plotted the prevalence of *P. falciparum* on the x-axis against the spatially lagged variables of hookworm, *T. trichiura* and *A. lumbricoides* on the y-axis. All points with values above the mean (i.e. above zero) were sites with high prevalence while all points with values below the mean (i.e. below zero) were sites with low prevalence. This holds true for the prevalence of *P. falciparum* plotted on the x-axis. The scatter plot was divided into four quadrants. The values of *P. falciparum* and hookworm, *T. trichiura* and *A.lumbricoides* that fell under the upper-right quadrant and the lower left quadrant were regarded as high-high (hotspots) and low-low (coldspots) values, respectively. We used the bivariate Moran’s I algorithm in Geoda software (https://geodacenter.github.io/) to implement this analysis.

### Spatial interpolation

To produce the prevalence surface of STH from the point referenced data, we used the inverse distance weighted kriging (interpolation) methods.[20] Prediction by kriging is based on the assumption that prevalence estimates at locations close together are correlated. Therefore, it allowed us to estimate the value of STH prevalence at any point from the observed data. We carried out the Kriging using ArcGIS version 10.3 software (https://arcview-gis.software.informer.com/10.3/). Our findings are virtualised at implementation unit level (administrative level 2 for most of the SSA countries). By implementation unit, WHO is considering the administrative division at which the preventive chemotherapy related interventions are to be implemented. For most of countries in the African region, these areas correspond with the second administrative level, although many countries have opted for using health district division when this exists.

### Population-weighted prevalence by implementation unit

We calculated the population-weighted prevalence of malaria and STH for each implementation unit using the population density data obtained from the WorldPop database (https://www.worldpop.org/project/categories?id=14) and the prevalence surfaces. Using 2020 Population raster of individual countries in SSA from the WorldPop database, estimated 165 million people were in high-risk clusters of *P.falciparum*, hookworm and *A.lumbricoides*. For malaria-STH co-infection, about 73 million, 21 million and 118 million people were in high risk clusters of *P.falciparum* and hookworm; *P.falciparum* and *T. trichura; P.falciparum* and A. lumbricoides; respectively. In most countries, administrative level 2 is the geographical area over which a particular treatment strategy or intervention is applied; this could be a district, local government area, county, or province. There were a total of 5,935 administrative level 2 in the study areas reported in this paper.

### Bivariate spatial clustering

To identify the spatial clusters of an implementation unit, we used local Moran’s I algorithm in Geoda software (https://geodacenter.github.io/) to implement the bivariate local indicator of spatial association (LISA).[21] LISA is a spatial autocorrelation used to assess the influence of individual locations on the magnitude of other locations. It gives an indication of the extent of significant spatial clustering of similar values around that observation. We used LISA to identify the hotspots and coldspots, and areas endemic to malaria-STH co-infection across the study areas. We considered areas with high malaria-STH co-infection and alpha level ≤0.001 as statistically significant hotspots.

## Results

The spatial auto-correlation plot was a linear fit whose slope corresponds to Moran’s I and its values for *P. falciparum* against hookworm, *T. trichiura* and *A. lumbricoides* were 0.302, 0.054 and 0.118, p-value <0.05, respectively (Fig 1A, B, C). High *P. falciparum* and high hookworm (HW) co-infection were more prevalent in the West and Central African sub-regions compared to other sub-regions of SSA (Figure 2A). The countries with high prevalence of *P. falciparum* and high HW co-infections were Angola, Benin, Burundi, Cameroon, Central Africa Republic, Congo Brazzaville,Cote d’Ivoire, Democratic Republic of the Congo (DRC), Ghana, Guinea, Kenya, Liberia, Mali, Nigeria, Sierra Leone, South Sudan, Togo, and Uganda (Figure 2A). Within these countries with high prevalence of *P.falciparum* and HW co-infection, there were specific areas (second administrative unit) with statistically significant co-infection (p < 0.001) which can be considered as hotspots for the co-infection.

**Fig 1A:**
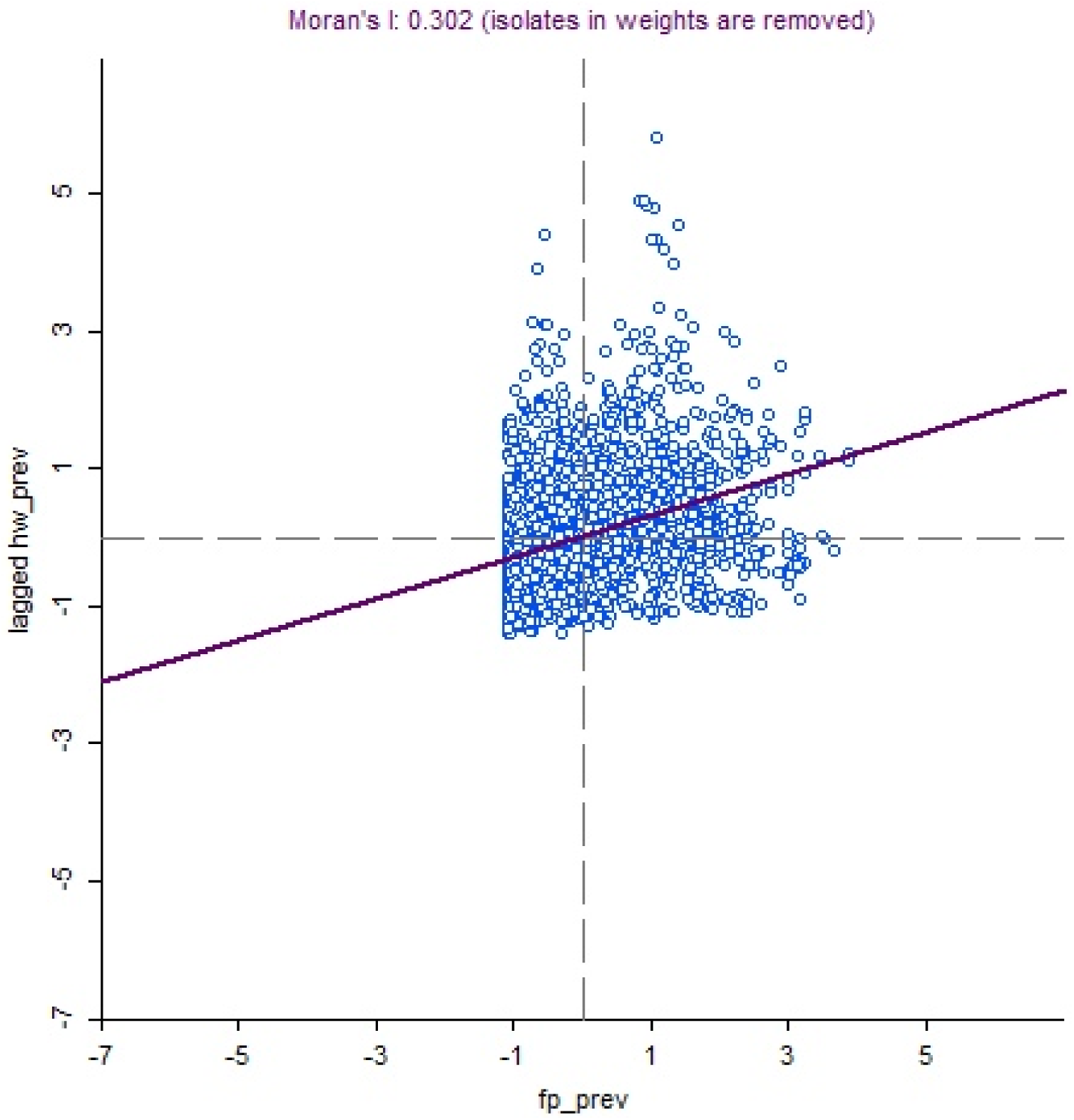
Moran’s I statistic plot showing spatial auto-correlation for *P.falciparum* vs hookworm co-infections in sub-Saharan Africa, 2000-2018.

**Fig 1B:**
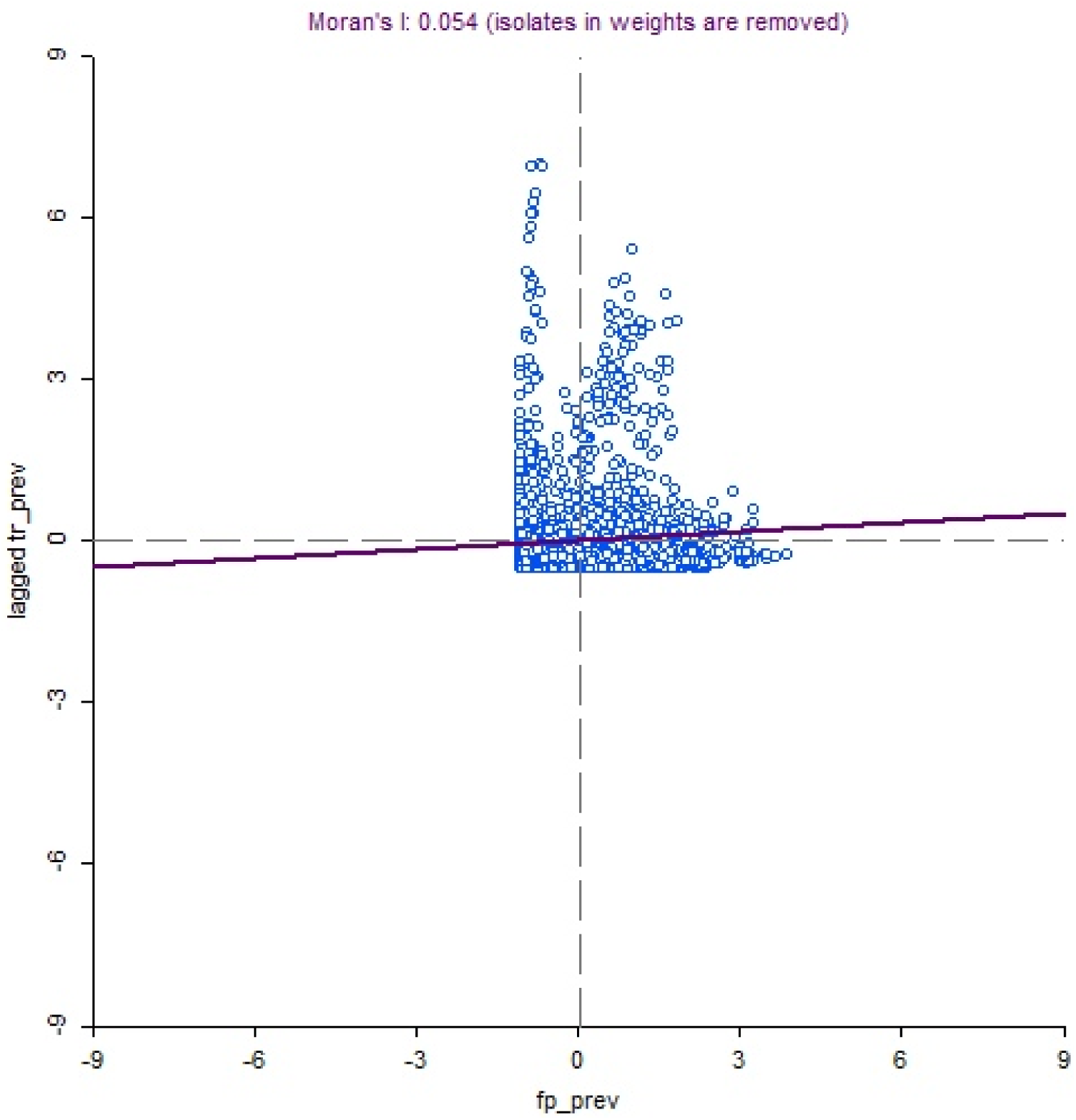
Moran’s I statistic plot showing spatial auto-correlation for *P.falciparum* vs T. *trichiura* co-infections in sub-Saharan Africa, 2000-2018.

**Fig 1C:**
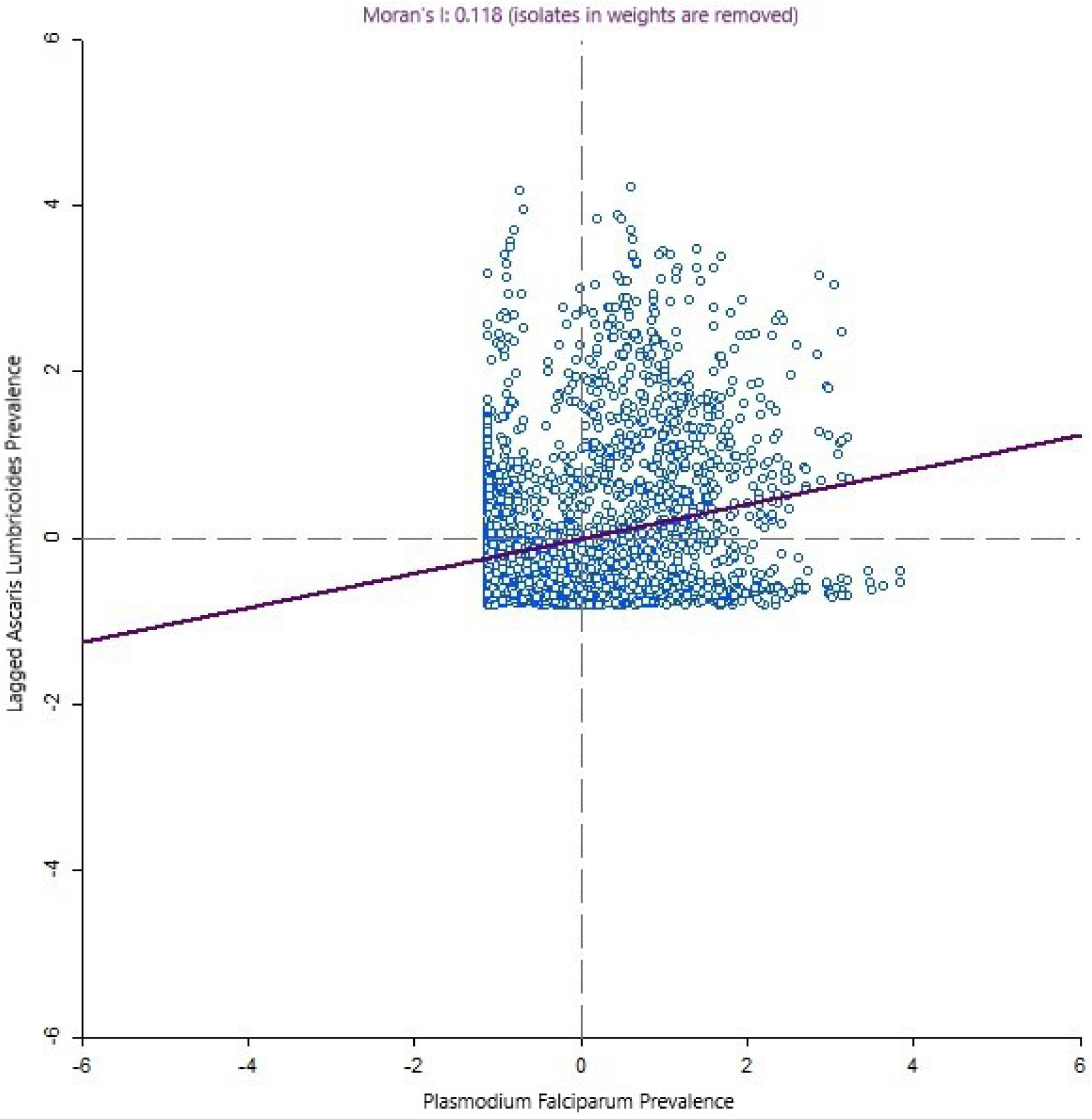
Moran’s I statistic plot showing spatial auto-correlation for *P.falciparum* vs *A.lumbricoides* co-infections in sub-Saharan Africa, 2000-2018.

**Fig 2A:**
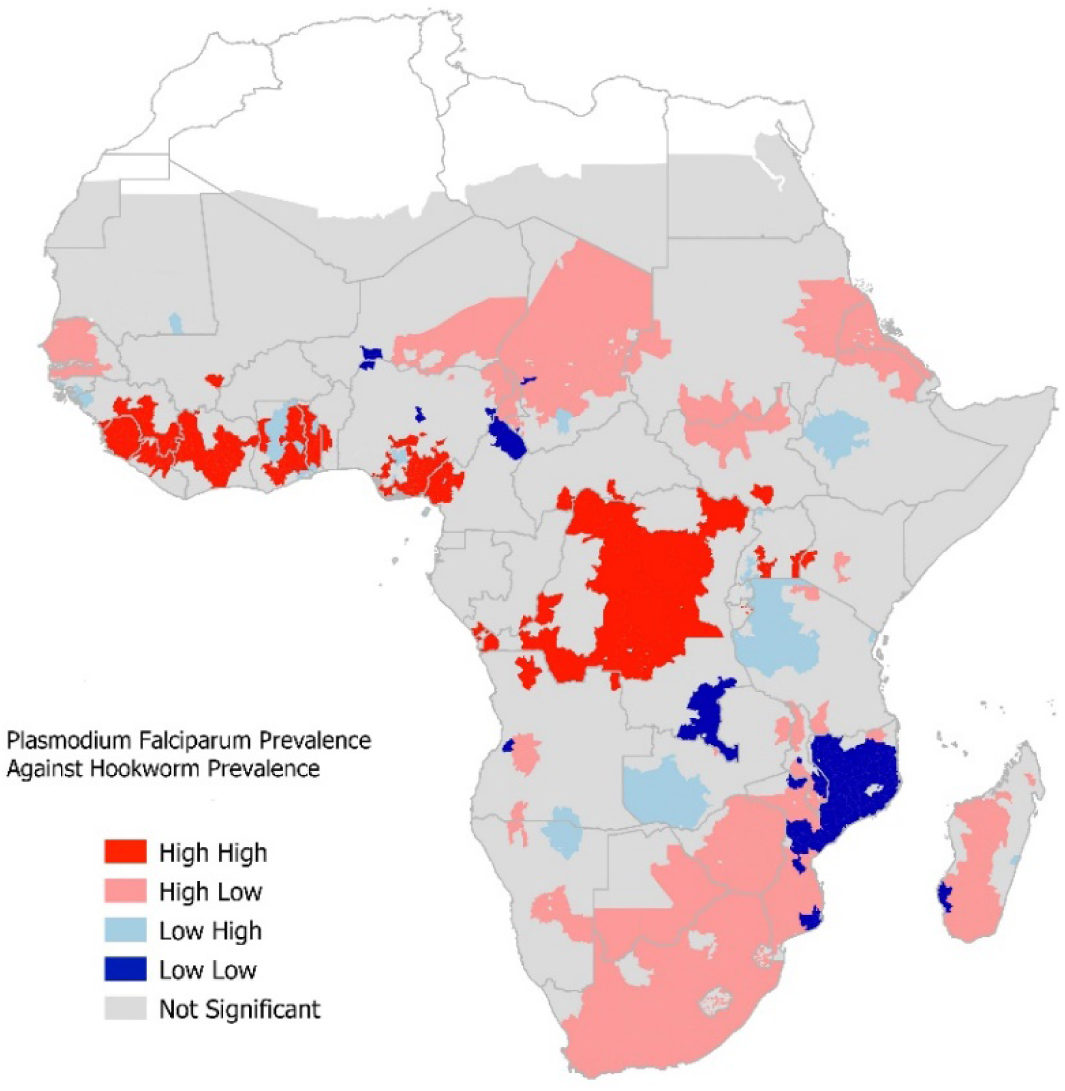
Local autocorrelation statistic interpretation of *P. falciparum* and hookworm co-infection, SSA, 2000-2018.

The second administrative units with statistically significant P.*falciparum* and HW co-infection (p<0.001) included north-west Angola; south-west Cameroon; west-coastal communities in Congo Brazzaville; south-central and east-central Cote d’Ivoire; and Kongo central province of DRC (Figure 2B). Other hotspots for co-infection with *P.falciparum* and HW were communities in western Kenya; south-western Liberia; eastern, southern and northern Sierra Leone; central and north-central Togo; districts in the Lake Victoria region and eastern Uganda (Figure 2B). In Nigeria, hotspots for co-infection with *P. falciparum* and HW co-infection included coastal communities in the southern and south-eastern parts of the country (Figure 2B, Table S1).

**Fig 2B:**
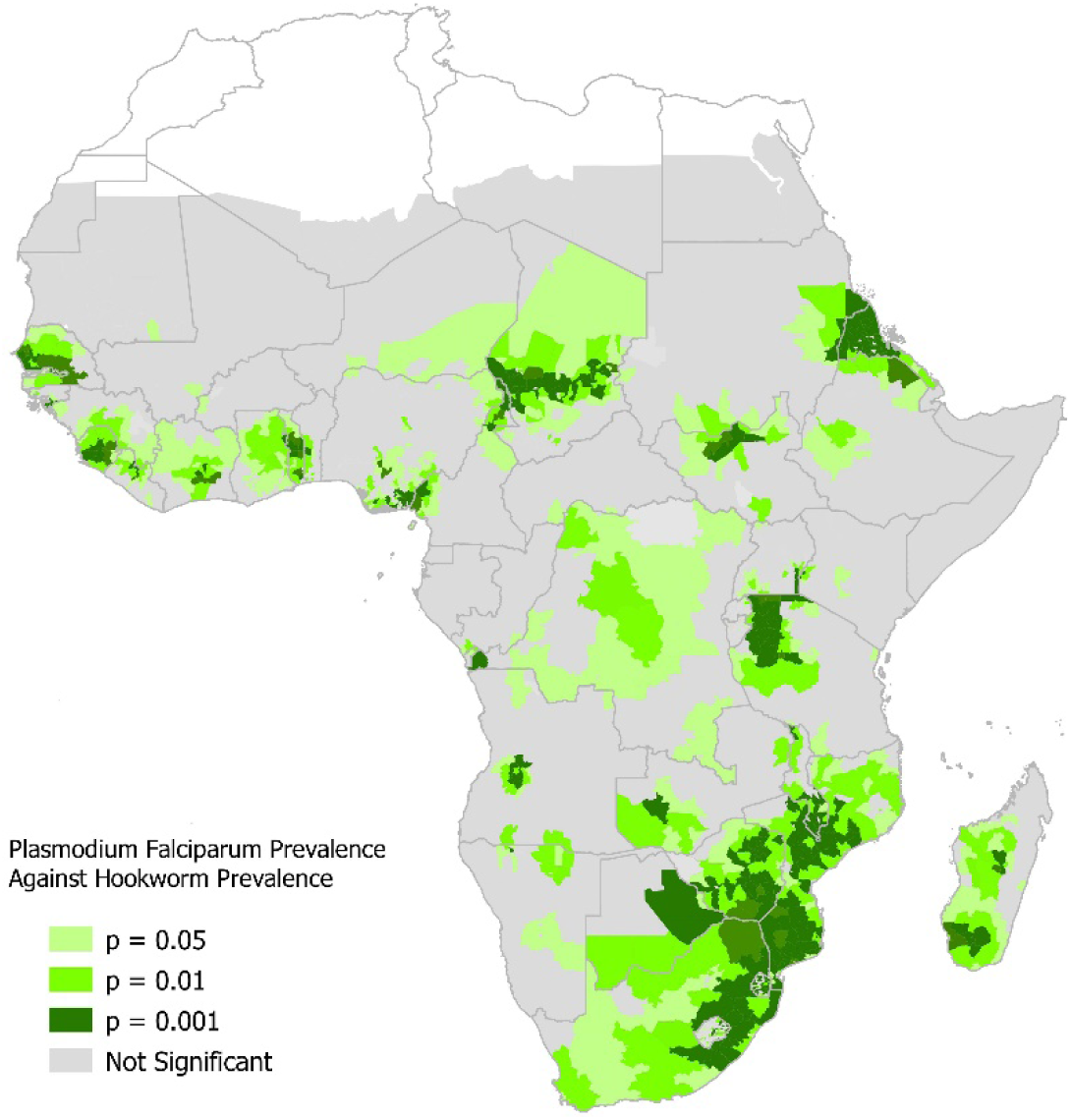
Probability that a given implementation unit is a hotspot for *P. falciparum* and hookworm co-infection in SSA, 2000 – 2018 *A.lumbricoides* co-infections in sub-Saharan Africa, 2000-2018.

High *P.falciparum* and high *T. trichiura* co-infection were more prevalent in the Central African sub-region compared to other sub-regions in SSA (Figure 3A). Angola, Cameroon, Congo Brazzaville, DRC, Equatorial Guinea, Gabon, and coastal communities of southern Nigeria had a high prevalence of *P. falciparum* and high *T. trichiura* co-infection (Figure 3A). Hotspots for *P. falciparum* and *T. trichiura* co-infections included communities around the coast in Angola and several communities in the central and southern Cameroon (Figure 3B). Other hotspots for co-infection with *P.falciparum* and *T. trichiura* were communities spread across all districts of Congo Brazzaville; all districts in the westernmost province of DRC; all regions in Equatorial Guinea and all districts except south-eastern region of Gabon (Figure 3B, Table S1).

**Fig 3A:**
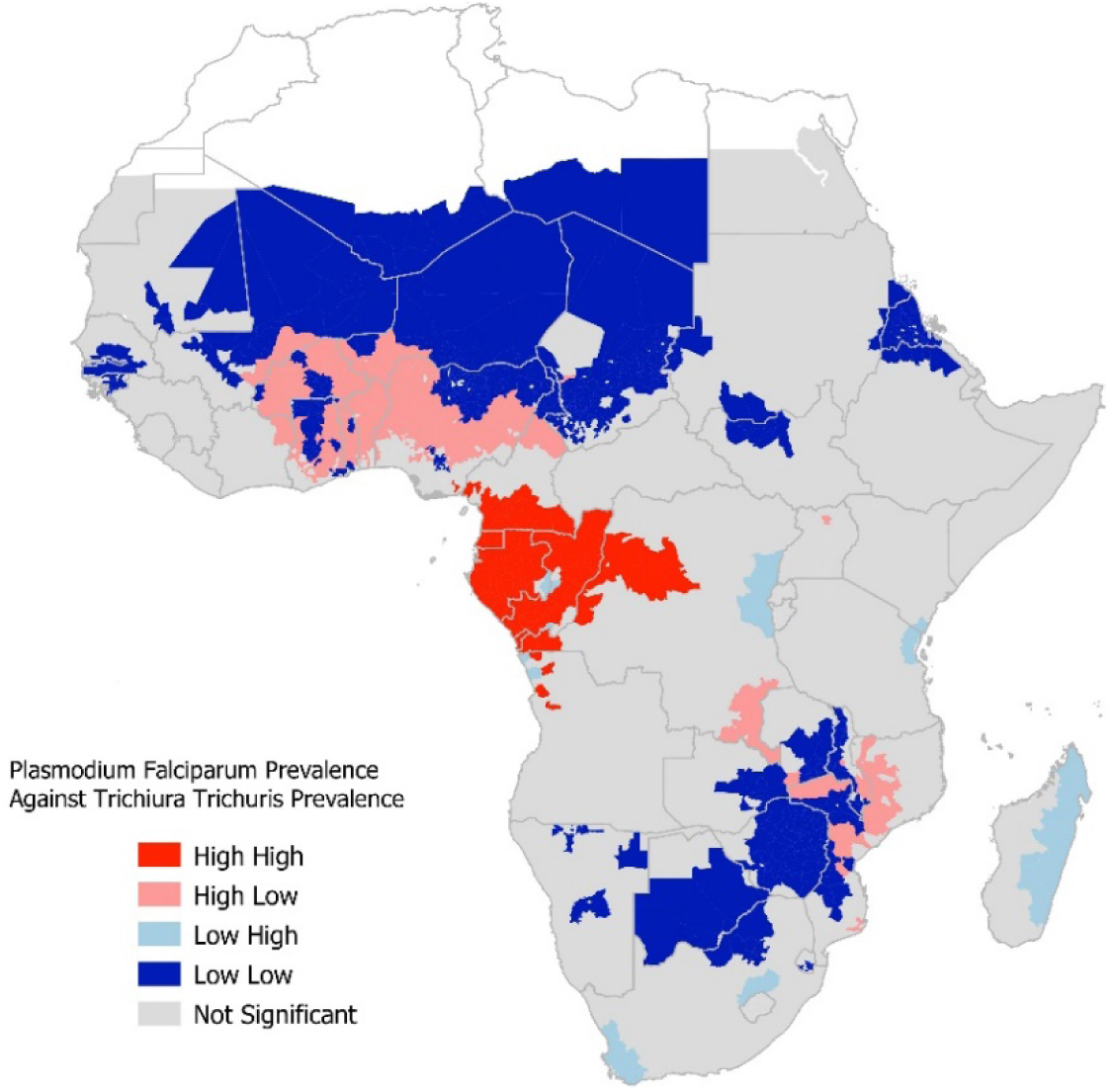
Local autocorrelation statistic interpretation of *P. falciparum* and *T. trichiura* co-infection, SSA, 2000-2018.

**Fig 3B:**
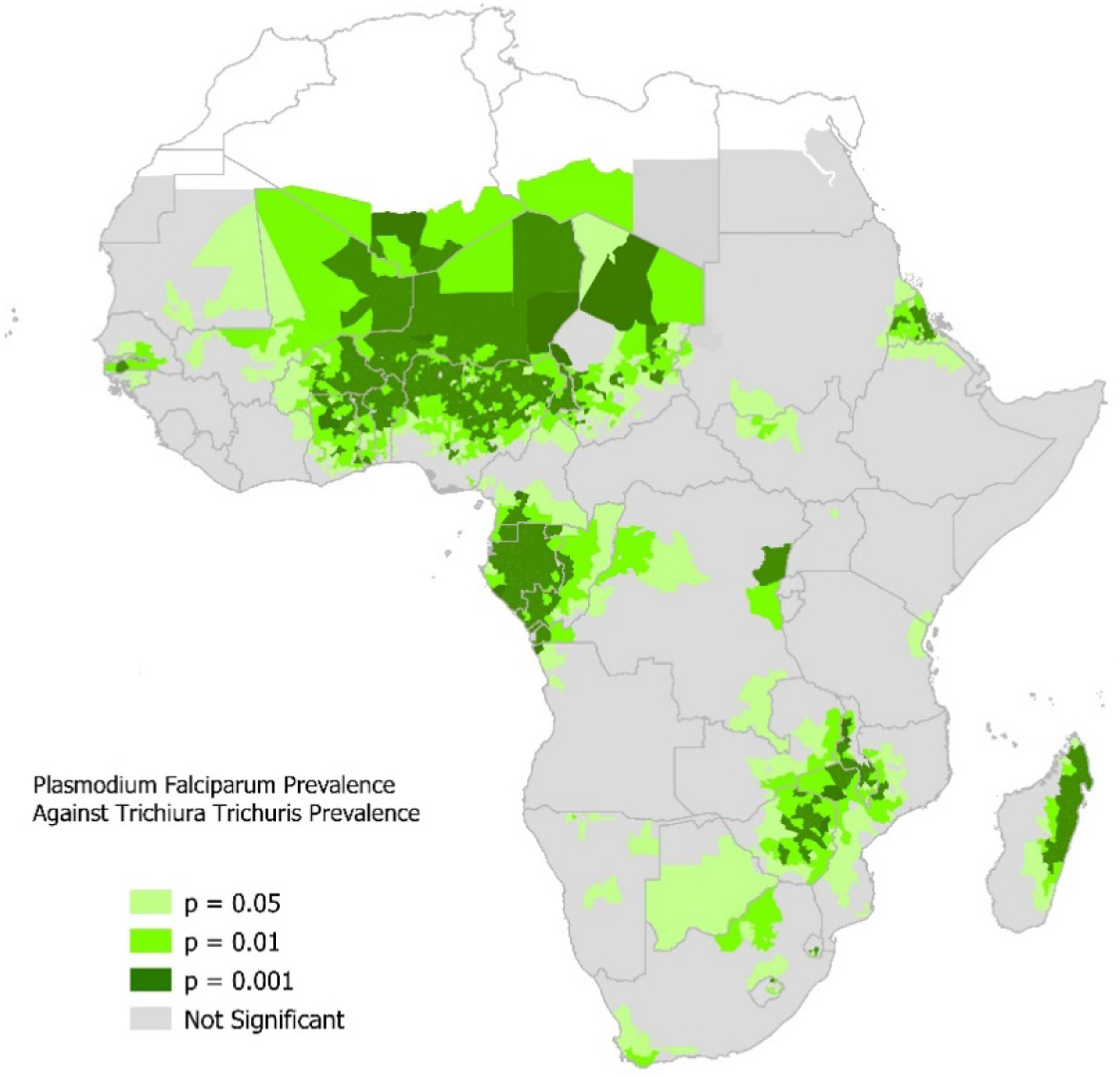
Probability that a given implementation unit is a hotspot for *P.falciparum* and *T.trichiura* co-infection, SSA, 2000 – 2018.

High *P.falciparum* and high *A. lumbricoides* (AS) co-infection was more prevalent in Central Africa and parts of West Africa (Figure 4A). The countries with high prevalence of *P.falciparum* and high *A. lumbricoides* co-infection were Angola, Burundi, Cameroon, Central African Republic, Congo Brazzaville, Cote d’Ivoire, DRC, Equatorial Guinea, Gabon, Kenya, Liberia, and Nigeria (Figure 3A). However, hotspots for *P.falciparum* and *A. lumbricoides* co-infection were identified in only seven countries: Angola, Congo Brazzaville, DRC, Equatorial Guinea, Gabon, Liberia, and Nigeria (Figure 4B).

**Figure 4A:**
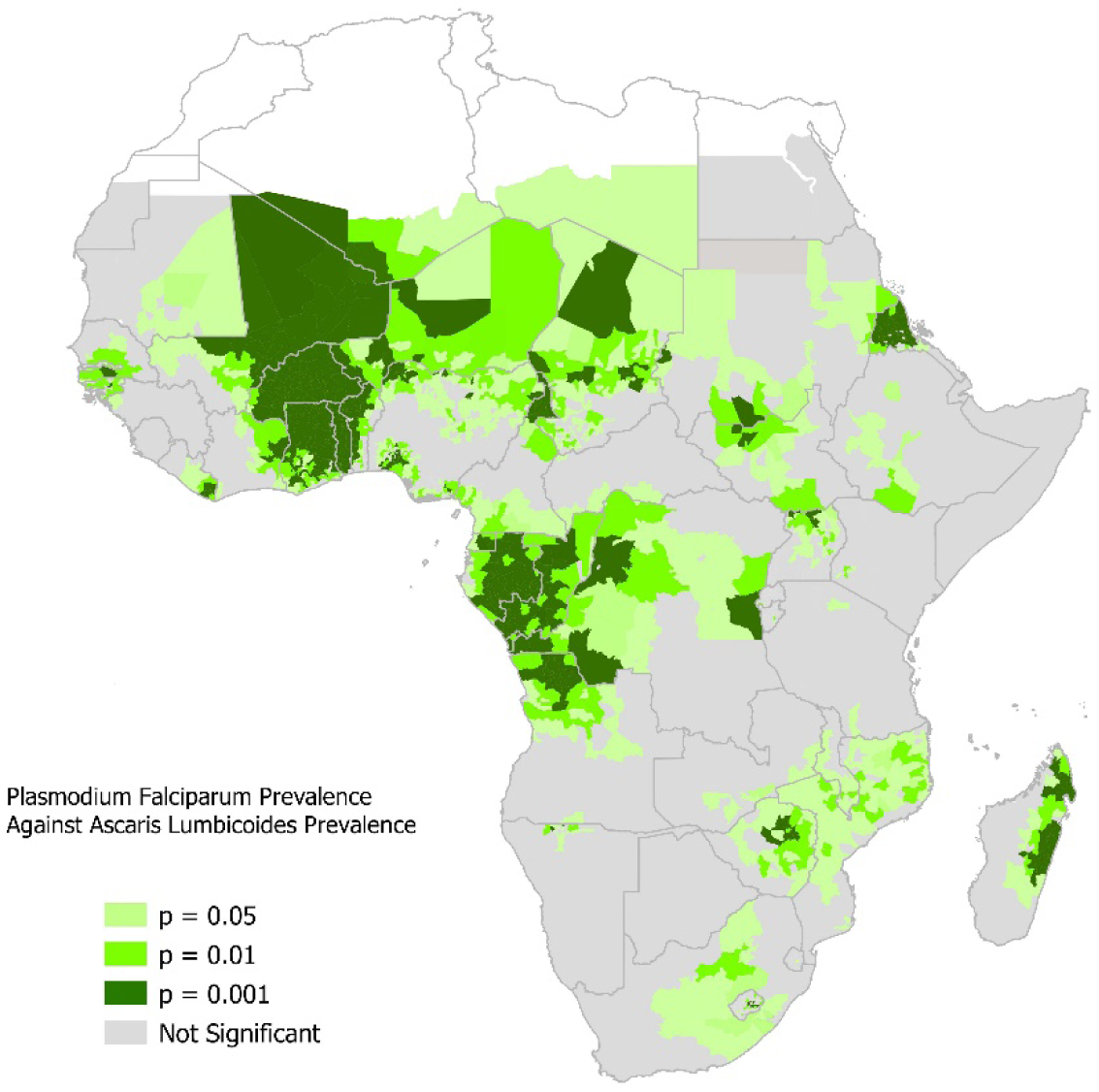
Local autocorrelation statistic interpretation of *P. falciparum* and *A. lumbricoides* co-infection, SSA, 2000-2018.

**Figure 4B:**
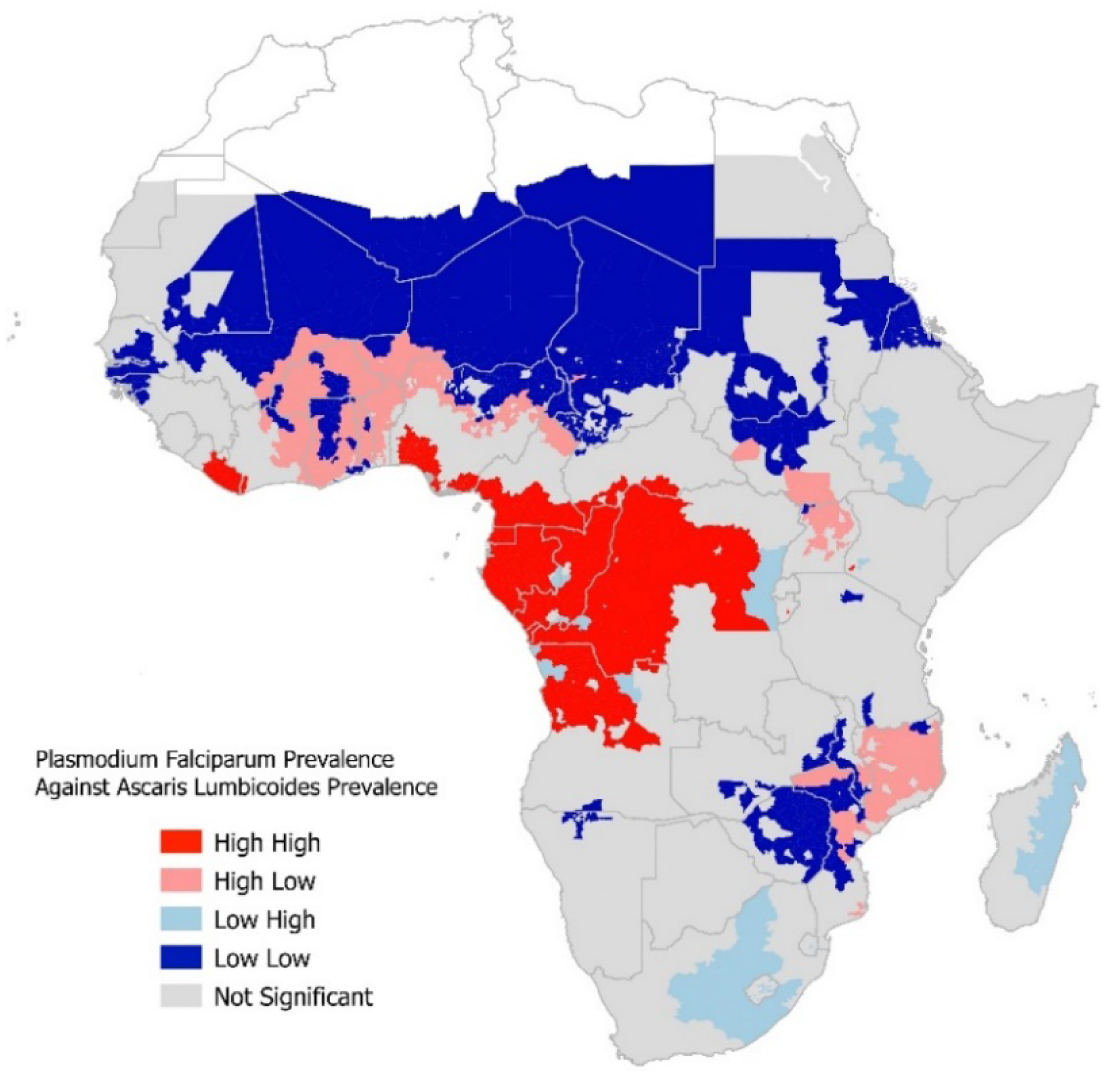
Probability that a given implementation unit is a hotspot for *P. falciparum* and *A. lumbricoides* co-infection, SSA, 2000 – 2018.

Hotspots for *P.falciparum* and *A. lumbricoides* in Angola included communities in the northeast and northern exclave of Angola, on the west (Atlantic) coast and north of the Congo River (Figure 4B). Other hotspots for *P.falciparum* and *A. lumbricoides* co-infections in SSA included north-western and south-western regions in DRC; all regions in Equatorial Guinea, the Littoral region and south-eastern counties of Liberia; and almost all regions in Gabon. In Nigeria, hotspots for *P.falciparum* and *A. lumbricoides* co-infections were in 13 states located along the coastal lines of the southern parts of the country (Figure 4B, Table S1).

## Discussion

Our analysis shows spatial variations in the estimates of prevalence of *P.falciparum*-STH co-infections and identified hotspots across many countries in SSA where the co-infections were high. Consistent with previous studies[3, 12, 22-24], our findings also highlight inter- and intra-country variations in the prevalence of malaria-helminth co-infections. Across Central and West Africa sub-regions, a high prevalence of *P.falciparum* and high hookworm co-infection was predominant, while the burden of *P. falciparum-A. lumbricoides* and *P.falciparum-T-trichiura* co-infections was higher in Central Africa than in neighbouring sub-regions. Although, recent reports have shown that a considerable gain has been made in the control of malaria and STH in the last two decades[1, 14], our findings suggest that a high burden of falciparum malaria and moderate-to-heavy infections for at least one STH species are still occurring in the southern part of West Africa and extending to Central Africa including Cameroon, Burundi and DRC. We observed similar patterns and trends in the hotspot distributions for *P. falciparum* and any of the STH. While programmatic implementation of preventive chemotherapy and seasonal malaria chemoprevention (SMC) have contributed to a significant reduction in the burden of STH and malaria in many countries, inherent challenges such as logistical burden, medication adherence, and incomplete coverage have been reported to be responsible for the persistently high burden of malaria and STH in the spatial areas identified in our study.[14, 15]

Taken together the geographical overlap and spatial co-existence of malaria and STH, favoured largely by environmental factors[1, 3], could be exploited to achieve effective control and elimination agendas through integration of the vertical programmes designed for malaria and STH into a more comprehensive and sustainable community-based paradigm, that includes other preventive care such as hand hygiene and water sanitation. The WHO 2030 NTD road map recommends concrete actions within integrated platforms for delivery of the interventions needed to improve the cost–effectiveness, coverage and geographical reach of these integrated programmes.[14] A successful integration of the vertical control programmes aligns well with the global trend towards integrated, non-disease specific approaches which have become an increasingly recognised strategy recommended by WHO.[25] However, limited evidence is currently available on the feasibility and effectiveness of integrating malaria with STH control at community level and evidence that this can be done effectively is needed.

Our study has some limitations. First, our study used secondary data obtained from two different sources covering a period from 2000–2018, which may not fully capture the epidemiological status of the burden of the co-infection during the period under review. Our analysis also showed some inconsistencies with findings of empirical studies [23, 26], especially for STH prevalence. For example, we observed high uncertainty around estimates in Central Africa and some parts of West Africa, which corresponds to areas with a high burden of infection. Lack of survey data is a plausible reason for this observation.[3] Additionally, a substantial proportion of the data we used for our analysis was collected prior to the commencement of mass preventive treatments with albendazole and SMC. Many countries have tailored the frequency of mass drug administration with preventive chemotherapy to their STH prevalence, but annual SMC implementation during the period of high malaria transmission is still being given as a single intervention.

The absence of survey data or inadequacy in definition of space-time contributes to the uncertainty observed in our finding and these may have also introduced bias leading to over-estimation or under-estimation of the prevalence of STH and malaria co-infection. Contributing further to the likely over or under-estimation of the burden of STH and malaria could be empirical treatments of malaria and STH outside the control programmes. Our findings could also be affected by the diagnostic approaches involving malaria rapid testing [27] and Kato-Katz method [28] used to generate the data, as these have been widely reported to be less sensitive than PCR, especially in low-transmission settings.

In conclusion, we have demonstrated wide spatial heterogeneity within the SSA and within countries, on the prevalence of malaria and STH confections. To consolidate on the encouraging progress made in the reduction of the prevalence of malaria and STH in some SSA countries, it is crucial that integrated control programmes target the regions with high-high burden of malaria and STH. Our study may also have critical implications for policy-making and resource allocations based on the needs of each region, as well as for the future of the implementation of integrated malaria-helminth control programmes. Nevertheless, it is important that our findings are confirmed by further empirical studies as this will provide the platform for research agenda that will lead to the establishment of the much-needed platform for the implementation of the integrated programmes that are cost-effective, and make optimum use of limited human resources frequently found in SSA.

## Data Availability

1. Malaria Atlas Project: https://malariaatlas.org/ 2. Expanded Special Project for the Elimination of Neglected Tropical Diseases: https://espen.afro.who.int/

https://malariaatlas.org/

## Acknowledgement

We thank Mike Thorn of the Malaria Atlas Project and Ismaela Abubakar for supporting the data curation for this study

## Author Contributions

MOA conceptualised the study. BS and JCO provided support in data curation and analysis. AA and OJ performed the statistical analysis of the datasets. MOA, OW, AA and OJ drafted different sections of the manuscript. BG revised the manuscript critically for important intellectual content. All authors reviewed and approved the final draft of the manuscript.

## Competing interests

None declared

## Funding

This work was implemented as part of a career development fellowship awarded to MOA, which is funded under the UK Research and Innovation Future Leaders Fellowship scheme (MR/S03286X/1). The funders had no role in the study design, data collection and analysis, decision to publish, or preparation of the manuscript.

## Notes

### Competing Interest Statement

The authors have declared no competing interest.

### Funding Statement

This study was implemented as part of a career development fellowship awarded to Muhammed Afolabi, which is funded under the UK Research and Innovation Future Leaders Fellowship scheme (MR/S03286X/1). The funders had no role in the study design, data collection and analysis, decision to publish, or preparation of the manuscript.

